# Assessing inertial measurement unit locations for freezing of gait detection and patient preference

**DOI:** 10.1101/2021.09.27.21264041

**Authors:** Johanna O’Day, Marissa Lee, Kirsten Seagers, Shannon Hoffman, Ava Jih-Schiff, Łukasz Kidziński, Scott Delp, Helen Bronte-Stewart

## Abstract

**Background:** Freezing of gait, a common symptom of Parkinson’s disease, presents as sporadic episodes in which an individual’s feet suddenly feel stuck to the ground. Inertial measurement units (IMUs) promise to enable at-home monitoring and personalization of therapy, but there is a lack of consensus on the number and location of IMUs for detecting freezing of gait. The purpose of this study was to assess IMU sets in the context of both freezing of gait detection performance and patient preference.

**Methods:** Sixteen people with Parkinson’s disease were surveyed about sensor preferences. Raw IMU data from seven people with Parkinson’s disease, wearing up to eleven sensors, were used to train convolutional neural networks to detect freezing of gait. Models trained with data from different sensor sets were assessed for technical performance; a best technical set and minimal IMU set were identified. Clinical utility was assessed by comparing model- and human-rater-determined percent time freezing and number of freezing events.

**Results:** The best technical set consisted of three IMUs (lumbar and both ankles, AUROC = 0.83), all of which were rated highly wearable. The minimal IMU set consisted of a single ankle IMU (AUROC = 0.80). Correlations between these models and human raters were good to excellent for percent time freezing (ICC = 0.93, 0.89) and number of freezing events (ICC = 0.95, 0.86) for the best technical set and minimal IMU set, respectively.

**Conclusions:** Several IMU sets consisting of three IMUs or fewer were highly rated for both technical performance and wearability, and more IMUs did not necessarily perform better in FOG detection. We openly share our data and software to further the development and adoption of a general, open-source model that uses raw signals and a standard sensor set for at-home monitoring of freezing of gait.

## BACKGROUND

Freezing of gait (FOG), a common symptom of Parkinson’s disease, is an intermittent inability to perform alternating stepping. FOG can occur when people attempt to initiate walking, turn, or navigate obstacles (1). It can lead to falls, injury, loss of independence, and decreased quality of life (2,3). Fifty percent of people with Parkinson’s disease experience FOG, and prevalence increases to 80% with disease progression (4,5).

Assessing FOG severity depends on subjective, qualitative tools like patient diaries, surveys, and clinical rating scales (6–9), or on observation by an experienced rater in a neurology clinic. Of these qualitative tools, the FOG Questionnaire, a six-question survey based on one’s experiences over the last week (10), is the recommended clinical assessment of FOG severity (11,12). While the FOG Questionnaire is more reliable than other clinical assessments (12), it does not offer granularity regarding FOG frequency and only quantifies length of freezes by patient recall. The gold standard of quantitatively measuring FOG is through post-hoc analysis of video-recorded gait by experienced raters (12). Features of the clinical setting, including the physical environment, on-medication status, and shift in attentional focus toward gait, limit FOG elicitation in patients who otherwise report FOG in their daily lives (13). The lack of a quantitative measure of FOG severity in a patient’s natural environment makes it difficult to track disease progression and tune therapy.

Advances in technology and machine learning methods enable objective FOG detection at low cost and in free-living settings (14,15). Inertial measurement units (IMUs) have high potential for capturing movement in any environment. Successful detection of FOG thus far with IMUs has required domain knowledge and years of development (16–19). In the past 15 years, more than 50 papers have reported FOG detection algorithms that use features devised by researchers or clinicians, including spectral analyses of accelerometer signals and gait parameters (16–18). A FOG detection model that does not use hand-engineered features relies less on domain knowledge and may be faster to develop. In addition, it may be more applicable across a variety of FOG presentations. Hand-engineered spectral features capture trembling, an increase in high-frequency leg movement in the relative absence of locomotion (20), that often accompanies freezing. However, there are other FOG presentations (21), and hand-engineered features may limit FOG identification to specific types of movement at specific body locations.

Studies have also probed a variety of sensor locations including the feet, shanks/ankles, knees, thighs, lower back, waist, chest, head, forearms and wrists (17,18). Despite the many locations explored, there is a lack of consensus about IMU location and quantity for the most accurate FOG detection. In addition, few studies investigate wearability and patient preferences, which may limit successful deployment. Recently, Davoudi *et al*. conducted a systematic analysis across accelerometer quantities and locations for physical activity recognition in older adults (22). This analysis informed the research community about sensor use in a general population across a variety of activities. A similar analysis among individuals with FOG could aid future research and practice in this population.

Monitoring FOG in natural environments can help us better understand its underlying mechanisms and inform therapy. It is important first to determine a standard IMU set that people with Parkinson’s disease will reliably wear and to develop a FOG detection algorithm that is accurate and robust. We address these needs using a combination of patient surveys, IMU measurements, and machine learning.

The aim of this study was to assess IMU sets in the context of both FOG detection performance and patient preference. This was supported by the development of an open-source framework to train neural networks that use raw IMU data to detect FOG without hand-engineered features. Assessments across sensor sets could aid future research protocol development and enable better adoption of IMU-based algorithms for FOG detection.

## METHODS

### Dataset

Our dataset consists of survey-reported IMU preferences and IMU walking data from a freeze-eliciting course.

Sixteen individuals with Parkinson’s disease, with 1 to 50 hours of prior in-clinic IMU experience, completed the survey (Appendix A). Participants were asked to extrapolate, from their in-clinic IMU experiences, the 12-hour wearability of individual IMUs and preferences for wearing various 3-, 2-, and single-IMU sets in a free-living setting. They were also asked to report motivations for their selections. IMU combinations included in the survey were selected from sets previously used to detect FOG in the literature (18).

Seven participants with Parkinson’s disease walked independently, without an assistive device, wearing IMUs (APDM Wearable Technologies, Inc., Portland, OR) through the turning and barrier course, specifically designed to elicit FOG. Each walking trial (walk) consisted of two ellipses and two figures of eight around tall barriers (23,24). Six IMUs were strapped on the tops of both feet, the lateral aspects of both shanks, and the lumbar (L5) and chest regions. Four of the seven participants wore a total of 11 IMUs, with five additional IMUs on the head, the posterior aspects of both wrists, and the lateral aspects of both thighs (Figure 1). Data were collected at 128 Hz.

Each participant provided 5 to 14 walks through the turning and barrier course across 2 to 6 clinic visits. Visits were separated by up to 44 months. In total there were 88.8 minutes of walking across 60 unique walks, with 211 unique FOG events resulting in 23.9% of the time freezing in the full dataset. Demographics and FOG frequency varied across participants (Table 1). A video of each walk was synchronized with the IMU system. Participants completed all trials off medication and off deep brain stimulation.

**Table 1.** Participant demographics and FOG frequency. Some values are reported as ranges because participants completed walks over multiple visits. (SD = standard deviation.)

| Participant | Gender | Disease Duration (y) | Number of Walks | % Time FOG (mean ± SD across walks) | Number of FOG Events (mean ± SD across walks) |
| --- | --- | --- | --- | --- | --- |
| 1 | M | 5-7 | 7 | 19.8 ± 13.5 | 4.4 ± 2.6 |
| 2 | M | 9-11 | 5 | 12.3 ± 25.0 | 1.8 ± 3.5 |
| 3 | F | 11-12 | 9 | 23.7 ± 11.1 | 4.2 ± 2.5 |
| 4 | F | 10-11 | 6 | 53.7 ± 19.8 | 8.0 ± 2.0 |
| 5 | M | 8-9 | 9 | 9.9 ± 11.3 | 2.0 ± 2.2 |
| 6 | F | 12-15 | 14 | 5.3 ± 5.8 | 1.4 ± 1.6 |
| 7 | M | 10-11 | 10 | 28.3 ± 19.8 | 4.8 ± 3.2 |
| **Total** | **M: 4**<br>**F: 3** | **Min: 5**<br>**Max: 15** | **60** | **19.7 ± 19.9** | **3.5 ± 3.1** |

Survey and IMU walking trial participants were not from the same cohort. Participants provided informed consent to a protocol approved by the Stanford University Institutional Review Board and completed the survey anonymously via REDCap.

### Data Processing

One of two experienced raters identified start and end times of FOG events in the video recordings of the walking data. A FOG event was defined by loss of alternating stepping, complete cessation of forward motion, or trembling of the legs (25).

IMU data were downsampled to 64 Hz and split into windows 2 seconds in duration, comparable to window lengths used in previous studies (16). Each window was normalized to zero mean and unit variance. To simulate variation in sensor orientation, the 2-second windows used to train the models were augmented with random rotations about the individual IMU axes (26). A window overlap was calculated to yield approximately 10,000 2-second windows. Each window’s label, FOG or not FOG, was set to the majority human-rated label of that window.

### Model Development

Two-layer, one-dimensional convolutional neural networks were trained using the labeled 2-second windows (Figure 2). We used convolutional neural networks because they are suited to small time series datasets. Relative to long short-term memory networks, convolutional neural networks contain few learnable parameters, which reduces overfitting to the training dataset. Our neural network architecture was parameterized with the number and lengths of filters in the convolutional layers and the number of neurons in the dense layer. We tuned these hyperparameters using leave-one-subject-out cross-validation. Hyperparameters were selected to increase the area under the receiver operating characteristic (AUROC) of the set with the most data, the 6-IMU set, to minimize overfitting. Hyperparameters remained constant across networks in our subsequent analysis. The resulting two convolutional layers contained 16 filters with kernel lengths of 17. Outputs were passed through a max pooling layer with length 2.

**Figure 2.**
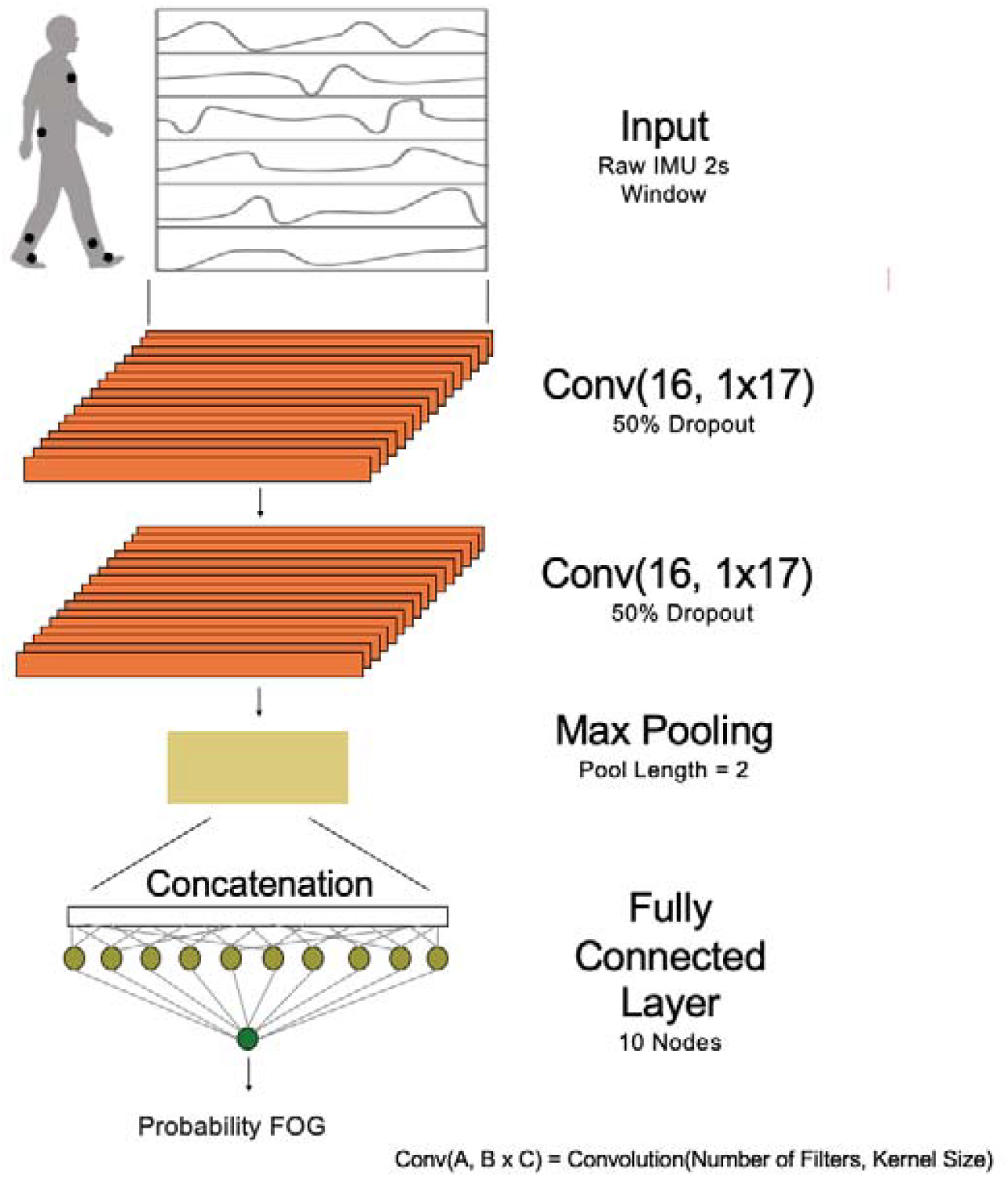
One-dimensional convolutional neural network model architecture. Two-layer, one-dimensional convolutional neural networks were trained using 2-second windows of raw inertial measurement unit (IMU) data. Each convolutional layer had 16 filters with a kernel length of 17. Weights were trained with 50% dropout. Convolutional layers were followed by a max pooling layer with a pool length of 2 and a 10-node fully-connected layer. The output of the model was the probability that the majority label for the 2-second window of raw IMU data was freezing of gait (FOG).

Concatenated data were passed through a 10-node fully-connected layer. The final node’s sigmoid activation function output the probability that the label for each 2-second window was FOG.

Models were trained with a batch size of 512, using the binary cross-entropy loss function and an Adam optimizer with a learning rate of 0.001. To avoid overfitting to certain participants or to the majority class during model training, we assigned equal weights to each class of each participant. Weights from each participant summed to 1, and weights from each of the two classes of each participant summed to 0.5. Dropout was implemented during training in the convolutional layers to reduce overfitting. Models were validated using leave-one-subject-out cross-validation. Early stopping to reduce overfitting was employed in each fold, using the loss of the left-out subject. Models were implemented using Keras (27) in TensorFlow 2.3.1 (28).

### IMU Sensor Set Experiments

We trained models with various IMU combinations to investigate how combinations performed relative to one another. We separately analyzed data from the four participants wearing 11 IMUs and the seven participants wearing 6 IMUs in order to compare results across sensor sets with data from equal numbers of participants.

Models with the full 11-IMU set and full 6-IMU set were trained to determine full sensor set performance. We also evaluated models with the 2- and 3-IMU sets from the survey as well as models with single IMUs. To account for stochasticity, each model was trained 30 times.

Model performance across IMU sets was evaluated using average held-out-set AUROC across folds. Two key IMU sets were identified: the best technical set and the minimal IMU set. The best technical set was defined as the IMU set that achieved the highest AUROC. The minimal IMU set was defined as the set with the fewest number of IMUs that still achieved an average AUROC within 5% of the best technical set (22). To account for IMU wearability and assess deployment feasibility, results from the best technical and minimal IMU sets were compared to results from the survey-preferred sets.

### Clinical Utility Assessment

Clinical utility was assessed on the best technical set and the minimal IMU set by computing intraclass correlations (ICCs) between model-identified and human-rater-labeled values of percent time FOG and number of FOG events for each walk. To translate our models’ FOG probability outputs into meaningful clinical metrics, probabilities were thresholded: examples with probabilities above a threshold were labeled FOG. To understand the upper bound of model performance, thresholds were selected to optimize each clinical outcome metric’s ICC with ground truth (i.e., one threshold for percent time FOG and another for number of FOG events). In practice, any threshold can be selected for a researcher’s or practitioner’s use case, trading off specificity for sensitivity.

Model-identified FOG periods that were one example apart were combined to a single FOG event, and short FOG periods that were just one example long were relabeled as non-FOG. The number of FOG events was calculated as the total number of non-FOG to FOG transitions in the walk. The percent time FOG was computed as the total duration of FOG events in the walk divided by the total duration of the walk, multiplied by 100. Clinical metric correlations between the model and a human rater were computed using ICCs. ICC estimates and their 95% confidence intervals were calculated using Pingouin (29) and were based on a single-rating, absolute-agreement, 1-way random-effects model. In accordance with Koo *et al*., (30), we used the following classification of reliability based on the ICC 95% confidence interval: < 0.50 poor, 0.50 to 0.75 moderate, 0.75 to 0.90 good, and > 0.90 excellent.

All analyses were performed using our custom framework, written in Python 3. Data and software can be found at *https://github.com/stanfordnmbl/imu-fog-detection*.

## RESULTS

### IMU Preferences

Individuals with Parkinson’s disease rated the wrist, ankle, and lumbar sensors as most wearable and the head and thigh sensors as least wearable (Figure 3a). Aside from the thigh and head sensors, more than 80% of respondents indicated a positive likelihood of wearing any individual sensor. In determining wearability, individuals reported perceived comfort and difficulty applying sensors as the most important decision factors (Figure 3b). IMU combinations with the greatest numbers of top rankings included: wrist and both ankles for a 3-sensor set, wrist and ankle or both ankles for a 2-sensor set, and wrist for a single-sensor set (Figure 3c).

**Figure 3.**
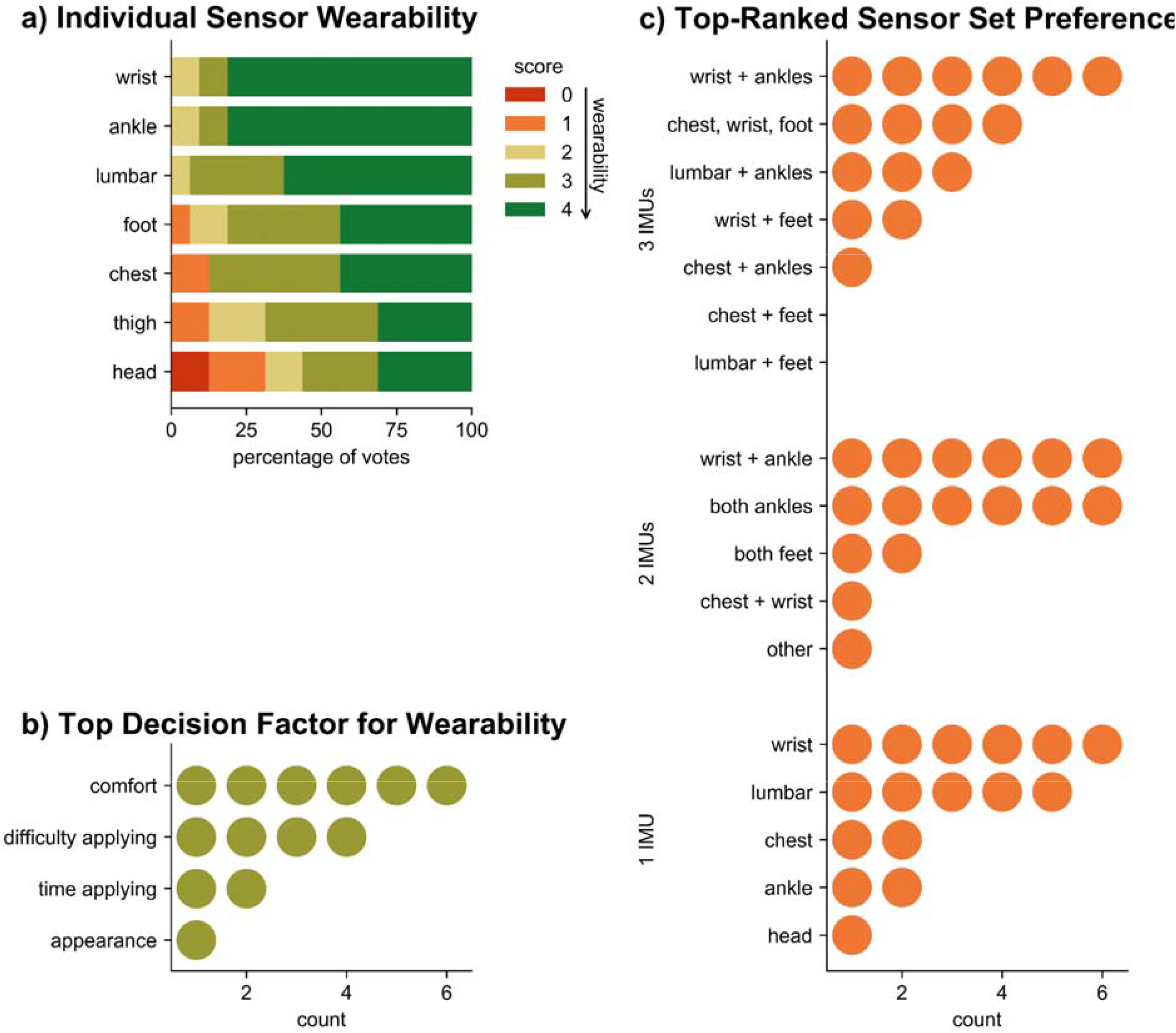
Survey results highlight patient preferences and wearability (n = 16). (a) Breakdown of individual sensor wearability scores. Scores ranged from 0 (definitely would NOT wear) to 4 (would happily wear, if asked). (b) Counts of top decision factor for determining wearability. Note, three participants did not answer the question associated with this, which involved ranking factors from least to most important. (c) Counts of top-ranked sensor set preferences for each of the 3-, 2-, and 1-IMU sets. Write-in sets (not offered as options) are indicated by “other.”

### IMU Sensor Set Experiments

A full-sensor model with data from all participants wearing 6 IMUs (chest, lumbar, both ankles, and both feet) estimated FOG from raw IMU signals with an AUROC of 0.80 (Figure 4a). Average precision (AP) of this model was 0.64, 2.5 times the positive predictive value of 0.26 (Supplementary Figure 1). The best technical set consisted of three IMUs, at the lumbar region and both ankles (AUROC = 0.83, AP = 0.66). Based on the survey, this set was also the preferred 3-IMU set among subsets of the 6 IMUs worn by all participants. The highest-performing 2-IMU set consisted of sensors from both ankles (AUROC = 0.82, AP = 0.63). This set was also one of the two preferred 2-IMU sets. The highest-performing single IMU was the ankle IMU (AUROC = 0.80, AP = 0.61). The preferred lumbar IMU demonstrated worse performance (AUROC = 0.75, AP = 0.54). The minimal IMU set, predefined as the set with the fewest number of IMUs that achieved an average AUROC within 5% that of the best technical set, consisted of a single ankle sensor (3.9% from best technical).

**Figure 4.**
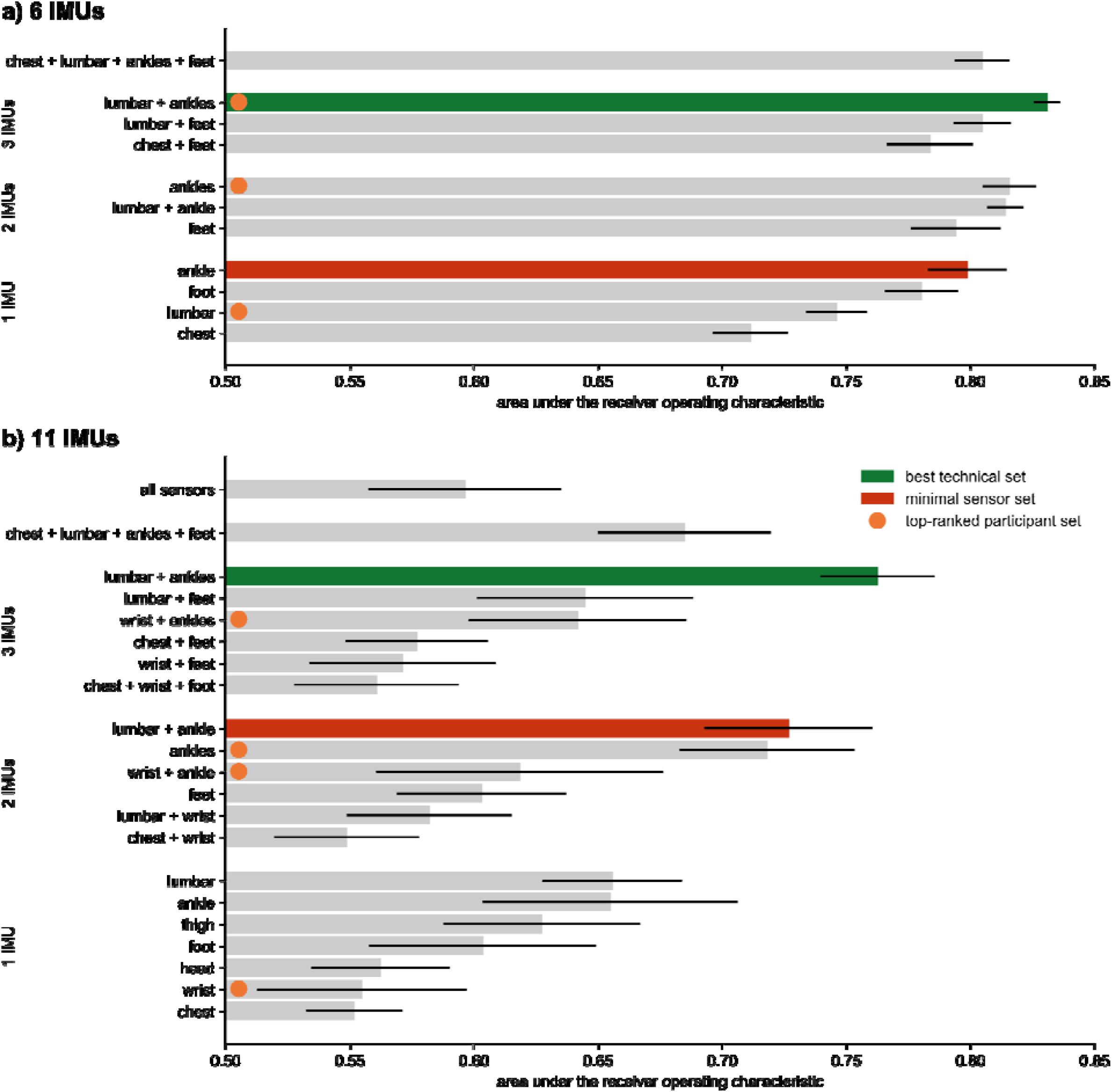
Test set AUROCs across subsets of the (a) 6-IMU and (b) 11-IMU sets. Best technical sets are indicated by green bars, and minimal IMU sets are indicated by red bars. Orange circles indicate the survey’s top-ranked sets for 1-, 2-, and 3-IMU subsets. Note, top-ranked sets differ across (a) and (b) because the 6-IMU set does not include wrist IMUs.

A full-sensor model with data from the subset of participants wearing 11 IMUs (head, chest, lumbar, both wrists, both thighs, both ankles, and both feet) estimated FOG with an AUROC of 0.60 (Figure 4b). The addition of the head, wrist, and thigh sensors to the 6-IMU set reported in Figure 4a did not change the best technical set (lumbar + ankles, AUROC = 0.76, AP = 0.35). The minimal IMU set with this subset of participants differed from the 6-IMU subset results and consisted of 2 sensors (lumbar + ankle, AUROC = 0.73, AP = 0.33, 4.7% from best technical) rather than a single ankle sensor. The highest-performing single IMUs in this data subset were the lumbar IMU (AUROC = 0.66, AP = 0.30, 14% from best technical) and ankle IMU (AUROC = 0.65, AP = 0.27, 14% from best technical).

### Clinical Utility Assessment

Interrater reliability between the best technical set (lumbar + ankles) and the individual human raters was good to excellent for percent time FOG (ICC = 0.93 [0.89, 0.96]) and number of FOG events (ICC = 0.95 [0.91, 0.97]) (Figure 5). Interrater reliability between the single ankle IMU and the individual human raters was slightly lower but still good to excellent for percent time FOG (ICC = 0.89 [0.82, 0.93]) and number of FOG events (ICC = 0.86 [0.78, 0.91]) (Supplementary Figure 2).

**Figure 5.**
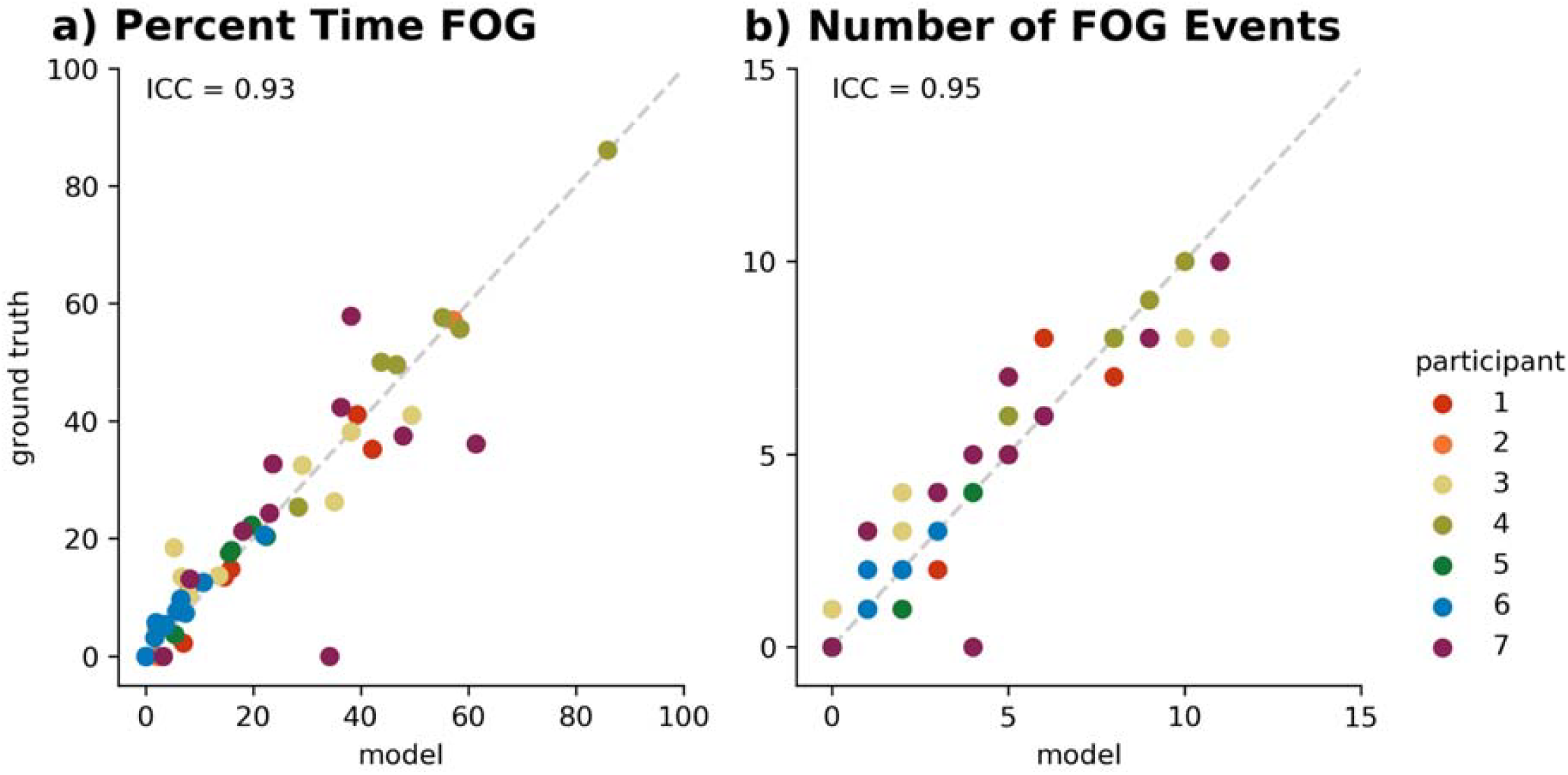
Clinically-relevant metrics from a model using the best technical set correlated with human ratings. Human raters’ ground truth vs model predictions from the best technical set (lumbar + ankles) for (a) percent time FOG and (b) number of FOG events. Intraclass correlations (ICCs) for the two metrics were 0.93 and 0.95, respectively. Each walk is depicted by a single datapoint. Data from individual participants are depicted by color.

## DISCUSSION

The goal of this study was to assess IMU sets in the context of both FOG detection performance and patient preference. Among pre-selected sensor sets, a best technical set and a minimal IMU set were identified. The best technical set consisted of only IMUs on the lumbar and both ankles, demonstrating that having more IMUs was not necessarily better. The minimal IMU set performed within 3.9% of the best technical set and used only a single ankle IMU. Both sets were highly rated for wearability and clinical metric performance. All models trained with data from seven subjects could detect FOG using raw IMU data (AUROC > 0.7). In conducting these analyses, we developed an open-source framework for identifying freezing of gait from raw time series data.

Based on raw IMU signals, the presented algorithm may be more applicable across various sensor locations and sets than an algorithm reliant on hand-engineered features. Further analysis should be conducted through a direct comparison of a hand-engineered spectral algorithm and our raw IMU algorithm.

Results from the 6-IMU set outperformed those from the 11-IMU set. In neural networks, more variation in the training data yields better model performance. Consequently, these results are expected, given that the 6-IMU set consisted of data from seven participants, while the 11-IMU set consisted of data from only four participants. Future model performance has the potential to increase further with more participants. Comparisons between different sensor subsets should not be made across the 6- and 11-IMU sets, and models should be evaluated by their relative, rather than absolute, performance within each set. Relative performances observed in the 6- and 11-IMU sets were largely consistent. The remainder of this discussion is focused on the 6-IMU set, which contained more training data and yielded better-performing models.

Within the 6-IMU set experiments, a 3-IMU subset consisting of the lumbar and two ankle IMUs achieved best technical performance, despite using hyperparameters tuned on the 6-IMU set. This demonstrates that more IMUs are not necessarily better for FOG detection, likely because additional IMUs can add more noise than signal. The 3-IMU best technical set produced clinically-relevant metrics that were in agreement with our human raters’ assessments (Figure 5). ICCs were improved compared to other algorithms and human raters (ICC = 0.73, 0.75, (31)) and between clinical raters (ICC = 0.76, 0.45, (32)) for percent time FOG and number of FOG events, respectively. In addition to detecting FOG, these three IMUs can provide further context relevant to a person’s gait and have been used in other algorithms (14,18,33). For example, the lumbar IMU can provide quantitative metrics of turning (34,35), and the bilateral ankle IMUs can provide metrics of regularity like arrhythmicity (23,24,36), both of which are relevant to clinical care.

Of note, a single ankle IMU performed at near-peak FOG detection performance. Despite ranked preference for a wrist sensor (Figure 3c), survey participants indicated equal individual wearability between the wrist and ankle sensors (Figure 3a). This high technical performance and wearability opens the door for simple data collection from a single IMU. It also suggests positive adoption of an ankle sensor by individuals with Parkinson’s disease who want to monitor and track their condition and treatments with minimal instrumentation. Not all single sensors performed well. While the IMUs at the lumbar region, ankles, and feet were found in the best-performing sets, IMUs at the wrists and head were less useful to the models. The wrist IMU, though rated highly wearable, demonstrated poor performance (AUROC = 0.56). This finding may be limited by the four-participant dataset, as a recent convolutional neural network trained on wrist IMU data from eleven participants showed comparable performance to our best technical set (sensitivity = 0.83 and specificity = 0.88, (37)).

We hope these results empower patients, clinicians, and researchers trying to weigh FOG detection performance with other monitoring needs across sensor locations.

### Limitations and Future Work

There are several limitations to consider before implementing this framework to detect FOG in a free-living setting.

Our survey captured sensor location preferences from individuals with Parkinson’s disease who had previously used the sensors in the clinic. Thus, the sample was skewed toward participants who generally wanted to aid research. This bias may have influenced results. Moreover, participants’ preferences were based on extrapolations from their in-clinic experiences, consistent with previous surveys on sensor preferences in other clinical populations (38). Survey results should be validated in individuals with IMU experience in free-living settings.

While good model performance was demonstrated for multiple IMU sets, our dataset and models had several limitations. All walking data were collected in a clinical setting rather than in a home environment. However, the turning and barrier course is explicitly designed to mimic movement in at-home settings. Additionally, participants were those with known FOG who completed walks off medication and deep brain stimulation. Translating to the broader Parkinson’s disease population will require training models with more diverse datasets that include participants who do not experience FOG and who are under different treatment conditions. Additionally, models were trained to detect one type of FOG, observed in continuous walking, and were not trained to identify FOG associated with gait initiation. For real-world application, a larger training dataset containing both FOG behaviors should be used.

Model training and assessment was limited by our small dataset. The observed decrease in model performance from a model trained with data from seven participants to a model trained with data from four participants suggests that a larger dataset will result in improved performance. Our models, using raw data and simple neural networks, underperform some previously-reported results from models that employ hand-engineered features (16,39,40). This is also likely due to our relatively small dataset, which limits the complexity of our models. A more complex model with more training data might perform comparably to a model with hand-engineered features. However, the objective of this study was to compare model performance across different sensor sets.

We present results from a small dataset and need to validate these findings on a larger, more diverse dataset. Sensor location preferences and model performance should be tested during real-life gait conditions on a larger cohort of participants with differing symptomatology and treatment.

## CONCLUSIONS

A framework was developed to train deep learning models on raw IMU data, enabling assessment of relative performances of sensor sets across the body. Experiments across sensor combinations and survey results demonstrated that three IMUs at the lumbar and ankles performed best in detecting FOG. A single ankle IMU performed within 3.9% of the best technical set. These two sensor sets agreed with patient wearability preferences and yielded clinically-relevant metrics, important for therapy personalization, that correlated well with human ratings. The dataset and framework (*https://github.com/stanfordnmbl/imu-fog-detection*) are intended to progress the community toward adopting a standard model and sensor set for FOG detection, an important step toward monitoring and personalization of care outside of the clinic.

## Data Availability

Sample code and data is available on github.

https://github.com/stanfordnmbl/imu-fog-detection

## List of abbreviations

(FOG): freezing of gait
(IMU): inertial measurement unit
(AUROC): area under the receiver operating characteristic
(ICC): intraclass correlation

## APPENDIX

### Movement Sensor Survey

#### Confidential

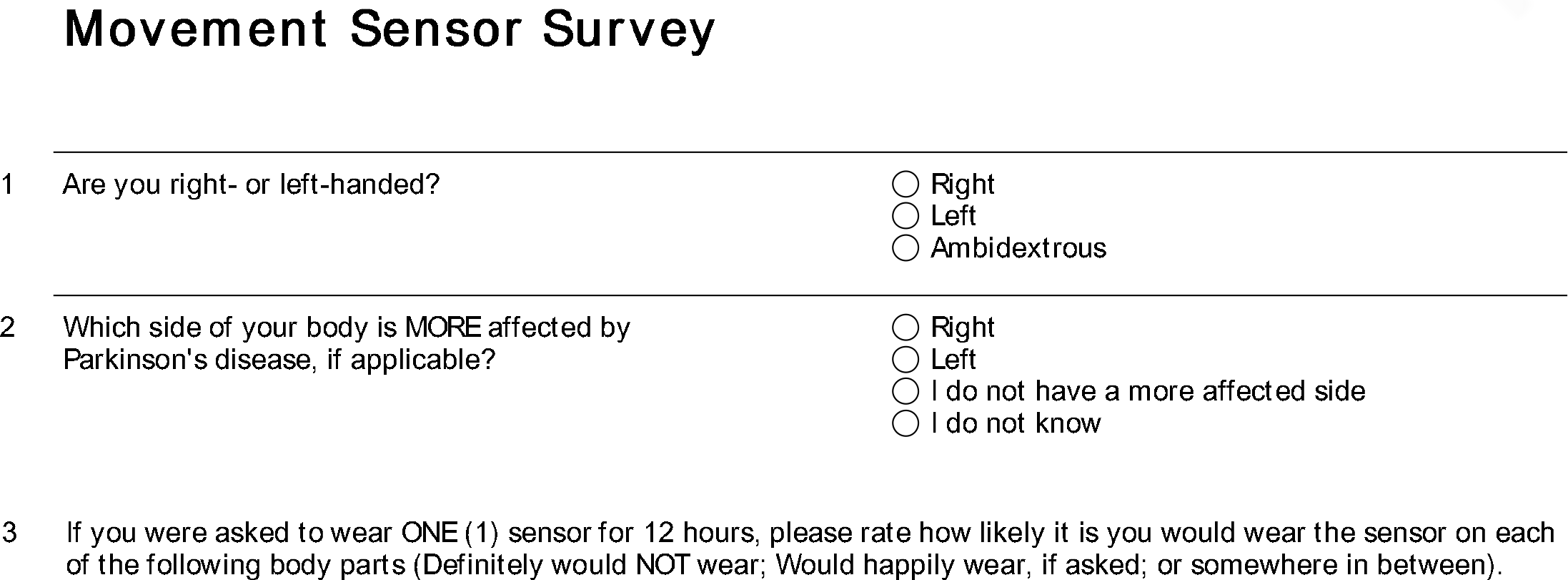

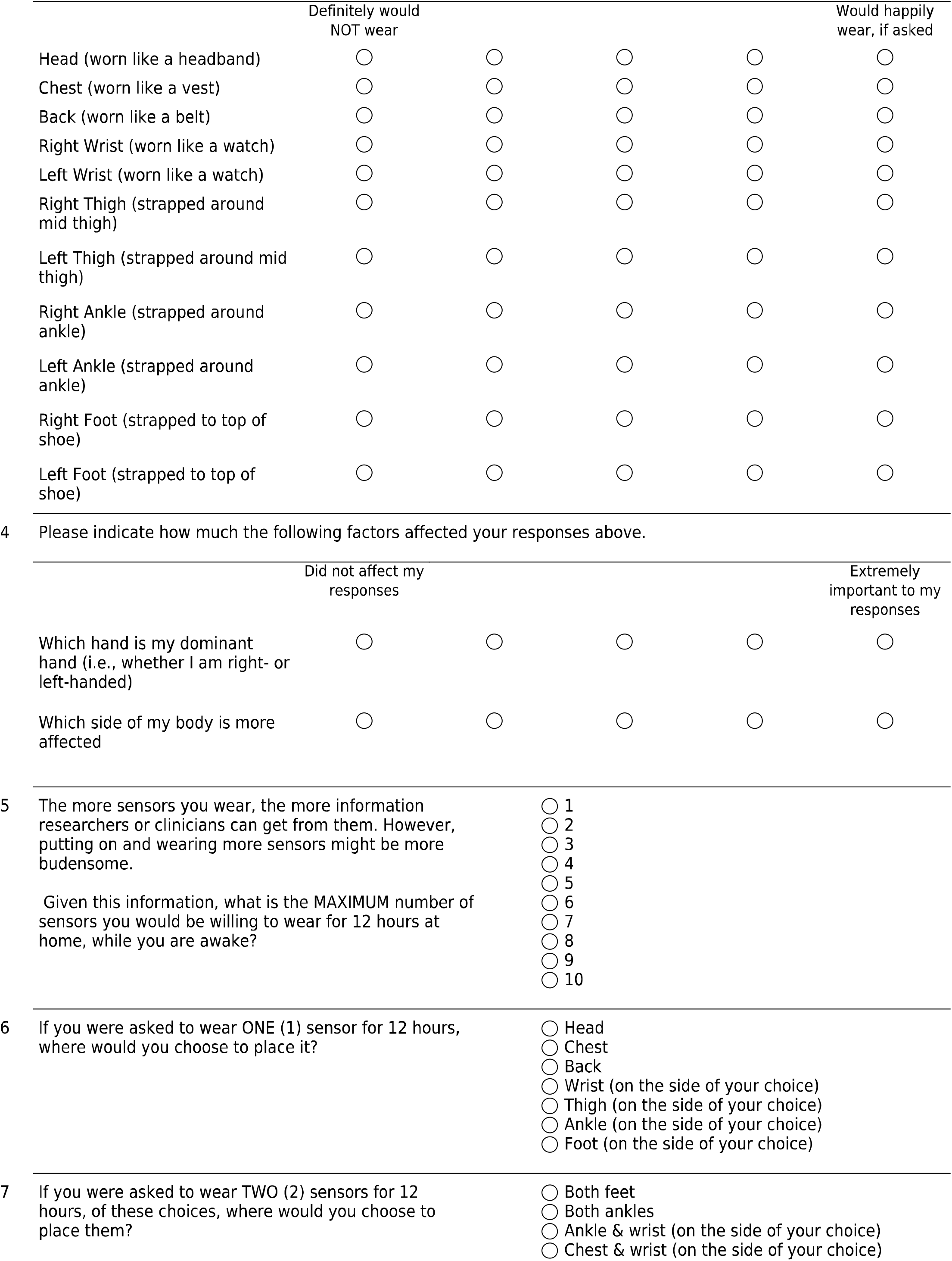

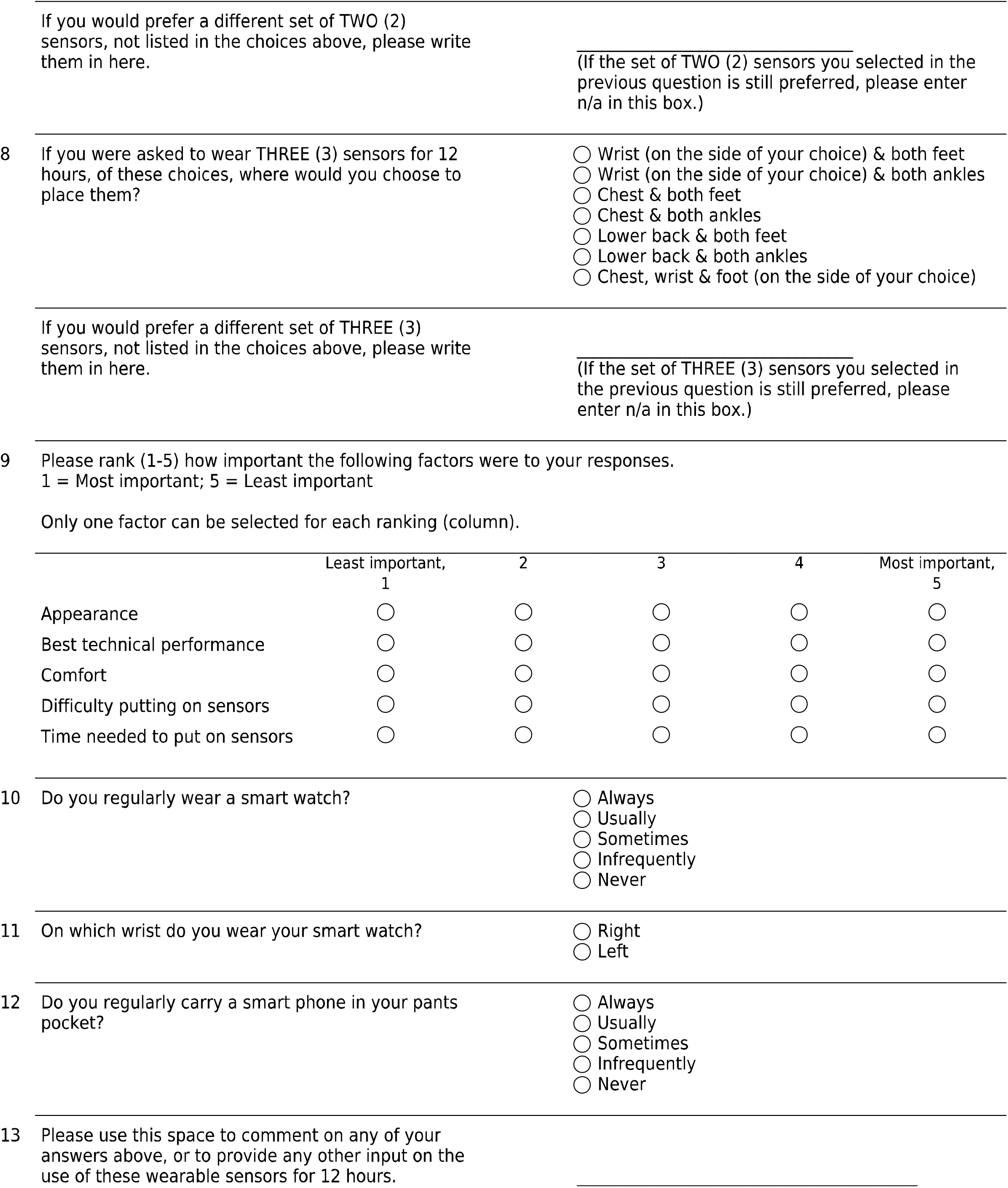

## DECLARATIONS

### Ethics approval and consent to participate

This study was conducted with the approval of Stanford Institutional Review Board.

### Availability of data and materials

The dataset and analysis scripts supporting the conclusions of this article is available on Github at https://github.com/stanfordnmbl/imu-fog-detection.

### Authorship contributions

- JO, ML, KS, LK, SD and HBS were responsible for the conceptualization of the project.
- JO, SH and HBS created the protocol and collected human subject walking data.
- JO and SH marked all freeze videos.
- SH created the survey (which was edited and reviewed by all first authors) and administered it via RedCap.
- AJS, SH and KS conducted survey data analysis.
- JO was responsible for initial processing of raw data (e.g. extracting from IMU sensors and aligning with raters’ labels).
- KS was responsible for pre-processing raw data before input to the neural network.
- ML, KS and JO wrote the software for data analysis and figures for the manuscript.
- ML cleaned code for the Github repository for optimal dissemination.
- SH, JO and HBS were in charge of the regulatory processes (e.g. with the Institutional Review Board) for study approval.
- JO, ML and KS wrote the original draft of the manuscript.
- HBS and SD supervised the project and acquired financial support.
- All authors were involved in interpretation of data and in writing, reviewing and editing the manuscript.

## Acknowledgements

We would like to thank the participants involved in the study as well Dr. Kevin Thomas for his support and expertise on the machine learning methodology.

